# A Multi-Granular Stacked Regression for Forecasting Long-Term Demand in Emergency Departments

**DOI:** 10.1101/2022.10.07.22280819

**Authors:** Charlotte James, Richard Wood, Rachel Denholm

## Abstract

**Background:** In the United Kingdom, Emergency Departments (EDs) are under significant pressure due to an ever-increasing number of attendances. Understanding how the capacity of other urgent care services and the health of a population may influence ED attendances is imperative for commissioners and policy makers to develop long-term strategies for reducing this pressure and improving quality and safety.

**Methods:** We developed a novel Multi-Granular Stacked Regression (MGSR) model using publicly available data to predict future mean monthly ED attendances within Clinical Commissioning Group regions in England. The MGSR combines measures of population health and health service capacity in other related settings. We assessed model performance using the R-squared statistic, measuring variance explained, and the Mean Absolute Percentage Error (MAPE), measuring forecasting accuracy. We used the MGSR to forecast ED demand over a 4-year period under hypothetical scenarios where service capacity is increased, or population health is improved.

**Results:** Measures of service capacity explain 41 ± 4% of the variance in monthly ED attendances and measures of population health explain 61 ± 25%. The MGSR leads to an overall improvement in performance, with an R-squared of 0.75 ± 0.03 and MAPE of 4% when forecasting mean monthly ED attendances per CCG. Using the MGSR to forecast long-term demand under different scenarios, we found improving population health would reduce peak ED attendances per CCG by approximately 600 per month after 2 years.

**Conclusions:** Combining models of population health and wider urgent care service capacity for predicting monthly ED attendances leads to an improved performance compared to each model individually. Policies designed to improve population health will reduce ED attendances and enhance quality and safety in the long-term.

## Background

In the United Kingdom, Emergency Department (ED) demand is increasing year-on-year [1]. In 2014 there were, on average, 61,318 ED attendances per day across the country; by 2019 this figure had risen to 70,230 attendances, an increase of 14.5% [2]. Given that the average cost of an ED attendance is £166 [3], this equates to an increase of over £500 million per year. Being able to forecast long-term ED demand is therefore important for strategic planning of services. Understanding the drivers of this increase in ED demand is also essential to policy makers to assess the potential impact of interventions for improving quality and safety by reducing demand and spending.

There exists a large volume of work that looks at forecasting demand on ED departments. Previous studies have mostly focussed on predicting short-term ED attendances [4-12] using past attendances [4,5,7,8, 10-14], calendar [5,7,9,10,14] and meteorological variables [5,9,14]. The most common techniques employed are Multiple Linear Regression [7,9,11,15], Autoregressive Integrated Moving Average (ARMIA) and variants [4,5,7,8,12,13], Exponential Smoothing [7,8,11] and, more recently, Machine Learning (ML) algorithms [5,8,10,12,13,16]. Cohort studies have found that measures of social deprivation and co-morbidities are predictive of ED attendances [17-19] and low General Practice (GP) attendance is associated with low ED attendance [18]. This suggests models used for forecasting need to incorporate measures of overall population health and access to other health services. As these variables change at different rates (GP availability changes daily, population health changes over a much longer time period), methods used in previous studies are not directly applicable.

Combining data at different scales falls under the paradigm of Granular Computing: a theoretical concept that recognises data contains different information when viewed at different resolutions [20]. Ensemble learning, a ML technique that combines the output from multiple independent algorithms into a unified framework, naturally lends itself to granular tasks. Multi-Granularity Ensemble Learning, where each algorithm in the ensemble is trained on data at a different scale, has been successfully applied to text [21,22] and image [23] classification and financial forecasting [24,25].

Here, we present a Multi-Granular Stacked Regression (MGSR) that uses measures of health service capacity with population and population health (including socioeconomic deprivation) to forecast long-term ED demand. Our model can be used by strategic planners and policy makers to determine how changes in service capacity or population health will impact long-term demand in ED departments.

## Methods

### Setting

This study uses publicly available data mapped to Clinical Commissioning Groups (CCGs) in England over the two-year period January 2018 to December 2019. We used this study period to avoid disruption from the COVID-19 pandemic. As NHS organisations, CCGs are part of a local network of healthcare providers: Primary Care, Secondary Care, Mental Health and Community Services. At the time of the study there were 81 CCGs in England with major EDs. Each CCG covers a certain geographical region, within which there are hospitals with ED Departments and GP surgeries. The CCG region also has a contract for 111, a 24-hour medical advice helpline, and a contract for Ambulance Services.

### Data

#### ED Attendances

ED attendance data was obtained from the A&E Attendances and Emergency Admissions dataset published by NHS England [26]. This data is released monthly and contains the total number of attendances at each NHS Trust for that month. NHS Trusts were mapped to CCGs. The outcome variable, ED attendances per CCG per month, was calculated as the sum of the attendances at each Trust for each month (see *Data Processing*).

#### GP capacity

Data on the number of GP appointments available in each CCG was obtained from the Appointments in General Practice dataset, published by NHS Digital [27]. This data is released monthly and is available stratified by CCG. The variable *count of appointments* corresponds to the daily number of GP appointments available. This variable was summed for each CCG to give a total number of appointments available per month as a measure of GP capacity.

#### 111 capacity

The NHS 111 minimum dataset was published monthly by NHS England up until March 2021 [28]. It contains detailed information on call handling and disposition after triage for all 111 contracts covering England. From this dataset, the variable *Calls offered* was used as a measure of the capacity of the 111 service. The daily number of calls offered for each contract was summed for each month before being stratified by CCG (see *Data Processing*).

#### Ambulance capacity

NHS England publish the Ambulance System Indicators (AmbSys) data monthly which contains information on calls made to the 11 ambulance service contracts covering England [29]. We used *Calls answered (A1)* as a measure of service capacity. The number of calls answered per month was summed for each contract before being stratified by CCG (see *Data Processing*).

#### Population

Estimates of the population of CCGs were obtained from the Office for National Statistics (ONS) [30]. Estimates of future population from 2019 to 2024 were also obtained to allow for forecasting of future ED demand.

#### Health Index

The Health Index is a measure of the health of the United Kingdom, consisting of three domains: *People, Places* and *Lives* [31]. Each of the three domains consist of a set of sub-domains. *People*, which quantifies health outcomes, is a combination of measures of mortality (e.g. life expectancy, avoidable deaths), morbidity (prevalence of long-term physical health conditions) and mental health conditions (e.g. prevalence of depression and anxiety, suicide rate). *Places* is a measure of wider determinants of health and environmental risk factors with components quantifying access to green space, quality of the local environment (e.g. air pollution, traffic noise), access to housing, access to services (e.g. distance to GPs and pharmacies) and levels of crime. *Lives* measures health-related behaviours and personal circumstances: physiological risk factors (e.g. diabetes, obesity); behavioural risk factors such as prevalence of alcohol and drug misuse; unemployment and working conditions (e.g. workplace safety); risk factors for children such as teenage pregnancy and child poverty; children and young people’s education; protective measures (e.g. cancer screening, vaccination coverage).

The overall index is a single indicator that can be compared over time and at different geographies. Values are relative to index values in 2015. It has a base of 100: values higher than 100 indicate better health than in 2015 and values lower than 100 indicate worse health. The Health Index is an experimental statistic developed by the ONS. Annual estimates are available stratified by CCG [32].

#### Data Processing

The curated dataset consisted of a single observation for each CCG and each month of the study period. For variables not reported at the level of CCG, data were aggregated (if reported at trust level) or stratified (if reported at contract level). Stratification was carried out according to population share: if a contract covers 3 CCGs, the amount of the variable assigned to each CCG will be proportional to the CCG’s population. When data was missing for a given CCG on a given month, the data point was removed from the data set. ED attendances, population and variables measuring the capacity of other services were scaled to units of per 10,000 people. As we are interested in forecasting future ED attendances, data for CCGs that have merged since 2018 were aggregated.

### Model Development

To forecast ED demand, we developed the MGSR to predict monthly ED attendances for each CCG. The model uses the capacity of other services, population and measures of population health as independent variables. We used a stacked architecture as measures of service capacity are available on a monthly basis, whereas population and measures of population health change annually. A stacked architecture allows for the predictions of independent models to be combined to produce a final prediction.

The MGSR consists of three separate models: two Level 0 models (Capacity Model and Population Health Model) and one Level 1 model, the Combined Model (Figure 1). The Capacity Model was trained on measures of service capacity as independent variables: the number of GP appointments available (GP capacity); the number of 111 calls offered (111 capacity); the number of ambulance calls answered (Ambulance capacity). The dependent variable was monthly ED attendances per CCG. The Population Health model had population and the three components of the Health Index (People, Places and Lives) as independent variables. The dependent variable was mean monthly ED attendances per year per CCG. The combined model takes the predicted values from the Capacity Model and the Population Health model to generate a final prediction for monthly ED attendances.

**Figure 1:**
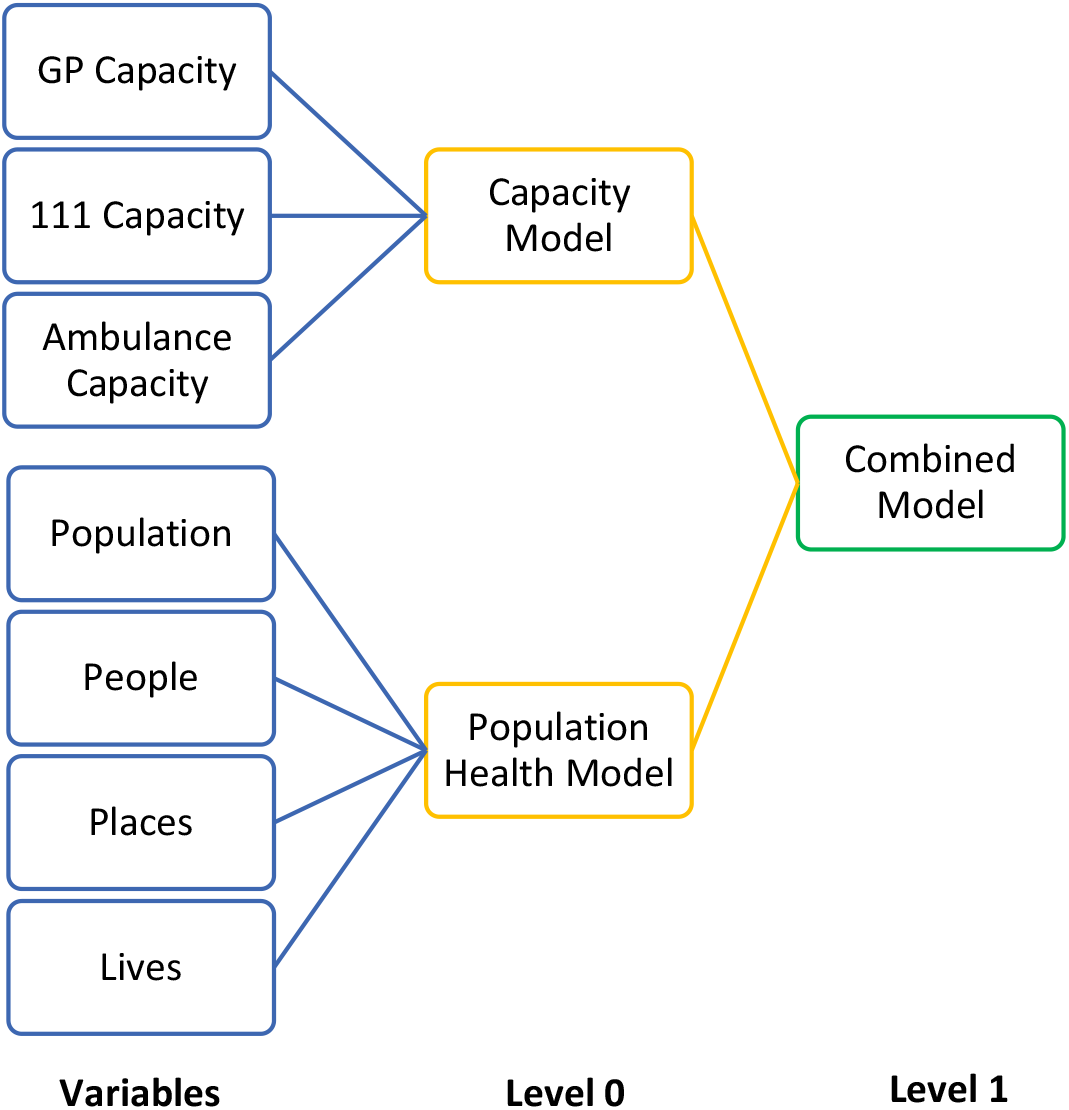
Schematic of the MGSR architecture.

For the Level 0 models we implemented a Random Forest (RF) algorithm [33] which is an example of an ensemble learning algorithm composed of individual decision trees. We used RF as it can handle non-linear relationships between independent and dependent variables, is known to be robust and has lower variance compared to a single decision tree. For the Combined model we implemented an Ordinary Least Squares (OLS) Regression due to the small number of variables and expected linearity between predicted values from the level 0 models and true values.

All models were implemented using Python’s sci-kit learn library [34], version 1.0.2. Repeated 5-fold cross-validation was used to assess individual model performance: data was split into a training set (80%) and validation set (20%) five times, such that each data point is in the validation set exactly once. This process was repeated 5 times with different random seeds, resulting in 25 different validation sets. At each fold, the model was fit to the training set and the validation set was used to assess performance. Final performance measures for each model were obtained by averaging over those for each validation set. For level-0 models, hyper-parameters controlling the number of trees and maximum tree depth were selected to maximise mean performance in the validation data while minimising model complexity [35].

To assess overall performance of the MGSR, at each fold the Capacity model was fit to 70% of the data (7/8 of the training set) and the level 1 model was fit to 10% of the data (1/8 of the training set). The Population Health model was fit independently to the Capacity model to ensure no data leakage between training and validation data [35]. The forecasting accuracy of the MGSR was assessed by training the model on data from 2018 only. This model was used to forecast both monthly ED attendances for each CCG, and mean monthly ED attendances per CCG, for 2019.

### Statistical Analysis

#### Model Evaluation

Individual model performance was evaluated using the coefficient of determination (R-squared), which measures the amount of variance in the dependant variable that is explained by the independent variables. The Mean Absolute Percentage Error (MAPE) was used to assess the forecasting accuracy of the MGSR.

#### Variable Importance

Variable importance was assessed for each model independently using the Gini Importance (GI)[36] for RF algorithms, and coefficient importance for OLS regression. Overall variable importance was determined using Permutation Importance (PI) [33]. Each variable in the data was randomly permuted sequentially, keeping all other variables constant. A variable’s PI was calculated as the difference between the model’s R-squared value without permutation and the R-squared when the variable is permuted. Repeated 5-fold cross-validation was used with 5 repeats. Final variable PI was obtained by averaging the difference in R-squared values for each fold and each repeat. A low PI suggests a variable has little influence on model performance whereas a high PI suggests an independent variable contains a lot of information about the dependent variable (monthly ED attendances).

### Forecasting Future Demand

For forecasting future demand, the MGSR was refit to the entire dataset. Population forecasts were obtained from the ONS. Data from 2019 was used as the baseline and ED attendances were forecast over a 4-year period. For each year, the population variable was replaced with forecasted population to obtain estimates of future ED demand in a ‘do nothing’ scenario (i.e., population health and service capacity remains as it was in 2019).

As we use data from 2019 as the baseline, theoretically the 4-year forecasting period represents 2020 – 2024. However, due to the impact of the COVID-19 pandemic on ED attendances and the capacity of other services, our forecasts will not be accurate for this period. We demonstrate the forecasting accuracy of the MGSR using pre-pandemic data (2018-2019) and separately use the model forecast future demand under different hypothetical scenarios had the pandemic not occurred.

#### Scenario Modelling

Future demand was forecasted under 4 different scenarios: 1) 111 capacity is increased by 10%; 2) GP capacity is increased by 10%; 3) Ambulance capacity is increased by 10%; 4) population health in the least healthy regions is improved. Together, these scenarios involve altering the independent variables of the MGSR either individually (scenarios 1-3) or in combination (scenario 4) to gain insight into how changes in service capacity or population health might impact future ED attendances. In scenarios 1-3, service capacity was immediately increased by 10%. For scenario 4, population health measures (*People, Places* and *Lives*) were increased in regions that scored below average in any of the measures in 2019. Specifically, if these variables were less than the baseline average, they were increased by 0.2 points per year until they reached the baseline average. By increasing levels of population health in the least healthy regions, rather than nationally, this scenario models how a reduction in health inequalities might impact future ED attendances.

## Results

### Data description

The curated dataset consisted of data from different publicly available sources (see Data). At the time of the analysis there were 81 CCGs in England with major EDs. Of these, 5 CCGs had incomplete data on ED attendances for 2018 and were removed from the analysis. This resulted in 76 CCGs and a maximum dataset size of 1824 datapoints. Datapoints were removed due to missing data in the following variables: Health Index (N=48); 111 capacity (N=158). This resulted in a final sample of 1618 data points containing 7 independent variables and representing 74 unique CCGs. Descriptive statistics for each independent variable are displayed in Table 1.

**Table 1:**
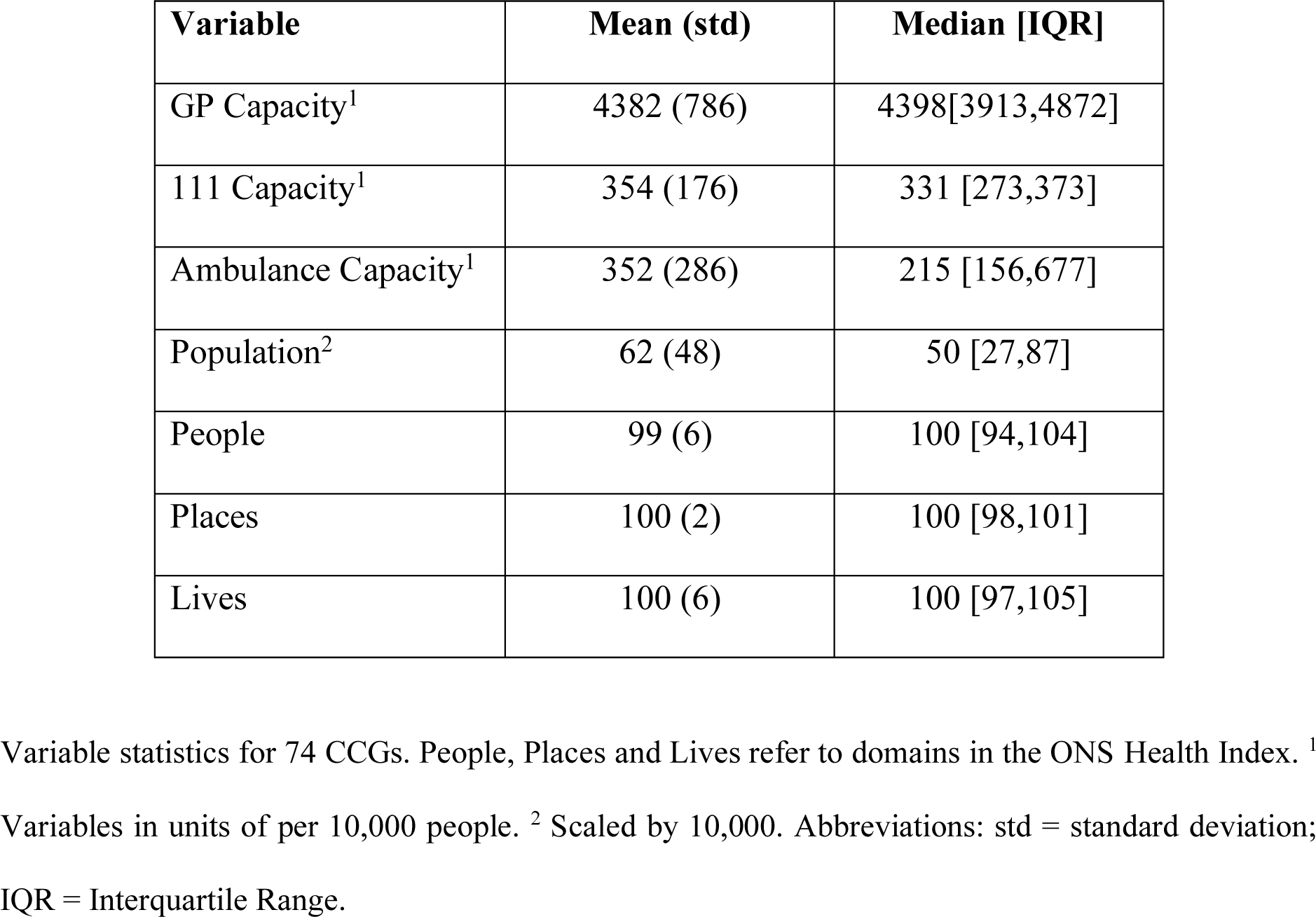
Independent variables used in the MGSR.

### Capacity Model

The RF fitted to measures of service capacity (GP, 111 and ambulance service) as independent variables was able to explain 41 ± 4% of the variance in monthly ED attendances. Of these variables, the most important was the capacity of the ambulance service (GI 0.65 ± 0.04) and the least important was GP capacity (GI 0.16 ± 0.02).

### Population Health Model

The RF fitted to population and measures of population health derived from the Health Index as independent variables was able to explain 61 ± 25% of the variance in monthly ED attendances. The most important variable was CCG population (GI 0.46 ± 0.19). Of the Health Index components, Lives was the most important (GI 0.33 ± 0.22) and People was least important (GI 0.08 ± 0.05).

### MGSR

Combining predicted mean monthly ED attendances (output of the Population Health model) and monthly ED attendances (output of the Capacity model) with an OLS Regression, the MGSR was able to explain 75 ± 3% of the variance in monthly ED attendances (Figure 2). The coefficient of the Capacity model was 0.37 ± 0.08 and the coefficient of the Population Health model was 0.75 ± 0.08 suggesting population and measures of population health are more predictive of ED attendances than the capacity of other healthcare services.

**Figure 2:**
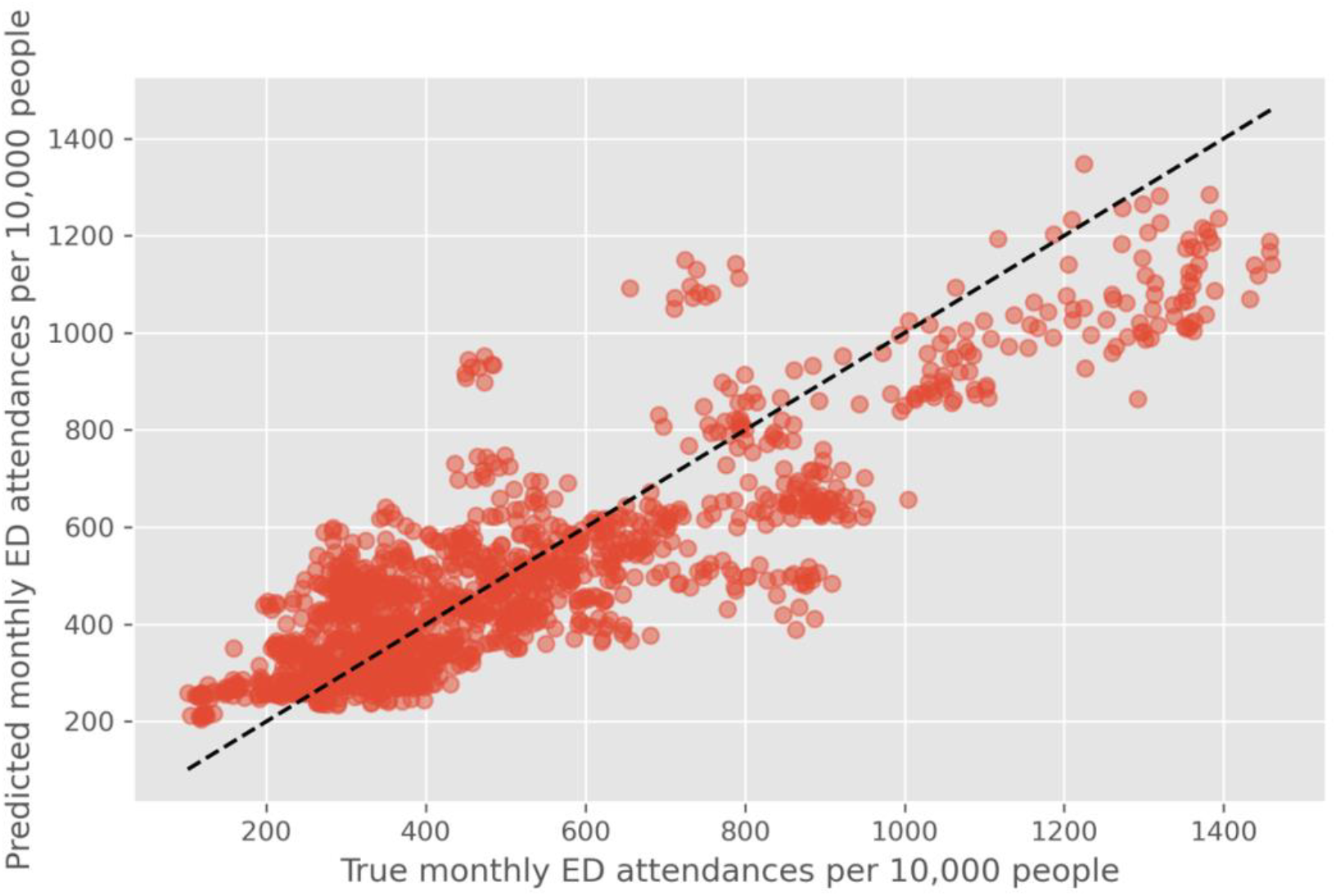
True ED attendances per 10,000 people (x-axis) versus predicted ED attendances per 10,000 people (y-axis) Black dashed line represents a perfect fit (R-squared = 1). Each point is a single CCG and a single month. Predicted values are averaged over 5 repeated 5-fold cross validations.

#### Forecasting Accuracy

Training the MGSR on data from 2018, the model was able to predict mean monthly ED attendances per CCG in 2019 with a MAPE of 0.04 (Figure 3). For all months except February, the predicted mean monthly ED attendances in 2019 (green dashed line, Figure 3) are less than the true mean monthly ED attendances (red dashed line). Over the winter period (December – February), the MGSR predicts mean monthly ED attendances per CCG most accurately, with an average error of 2%.

**Figure 3:**
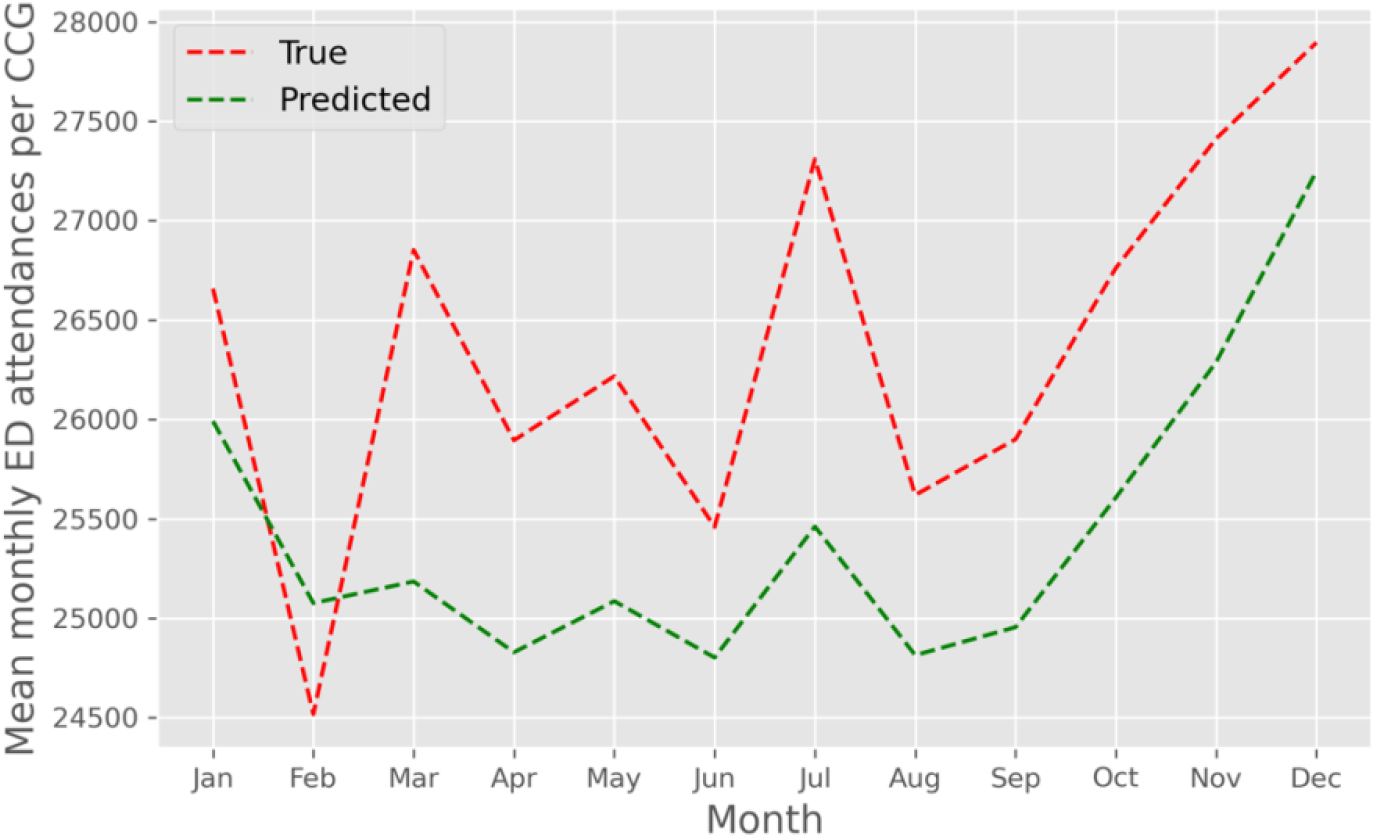
Forecasting accuracy of the MGSR. True mean monthly ED attendances per CCG (red dashed line) and predicted mean monthly ED attendances per CCG (green dashed line).

#### Variable Importance

We assessed overall variable importance by randomly permuting each variable individually and calculating the mean change in performance of the MGSR (Figure 4). The most important variable was population (PI 0.40 ± 0.08). Of the measures of service capacity, ambulance capacity is most important (PI 0.11 ± 0.04), followed by 111 capacity (0.05 ± 0.02) and GP capacity (PI 0.02 ± 0.01). Components of the Health Index (People, Places, Lives) are also important in predicting monthly ED attendances, with Lives providing the most information (PI 0.20 ± 0.04) followed by Places (PI 0.09 ± 0.03) and People (PI 0.06 ± 0.02).

**Figure 4:**
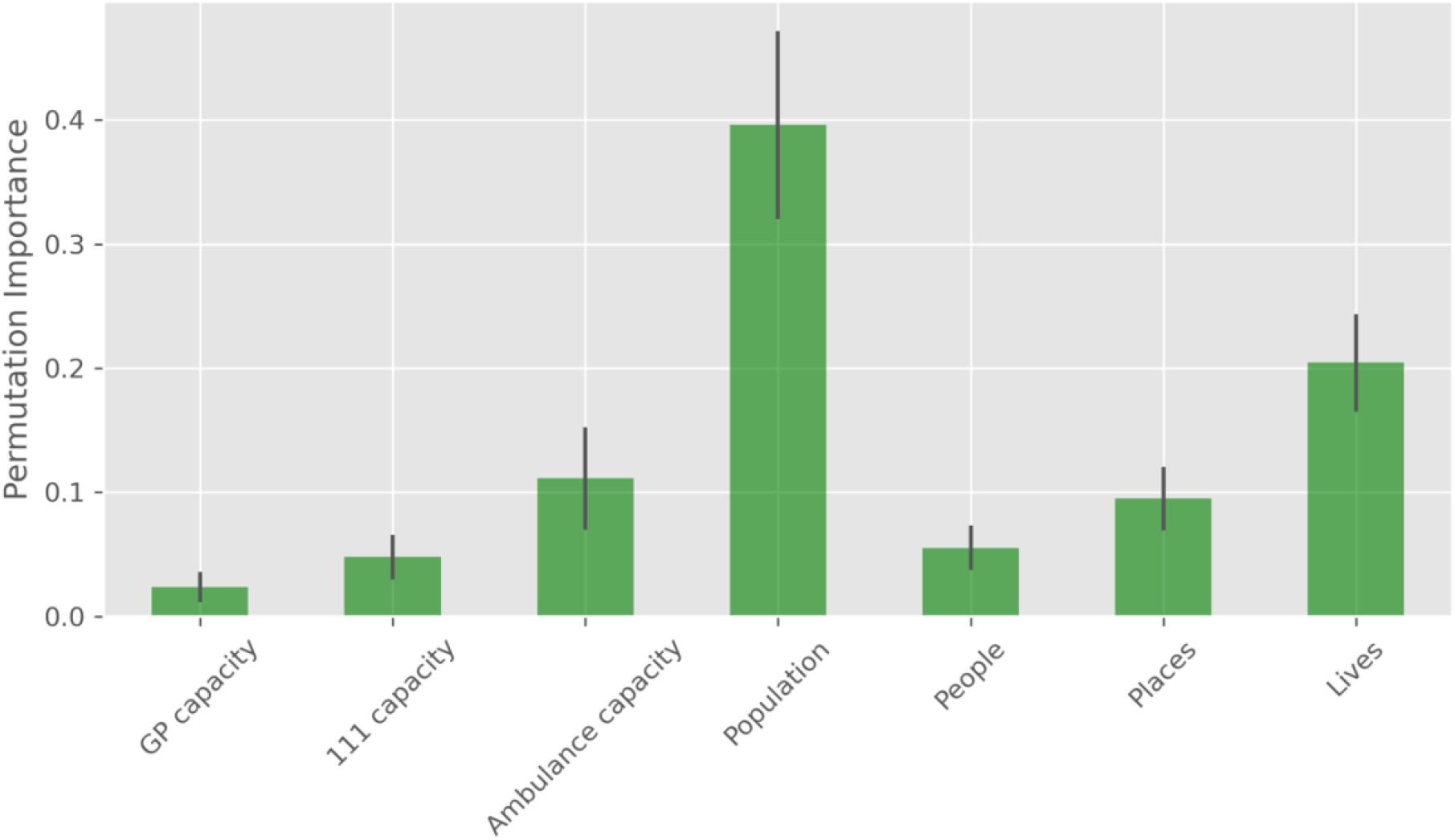
Permutation feature importance of the MGSR. Green bars represent mean importance from repeated 5-fold cross-validation. Black lines correspond to standard deviation in importance.

### Forecasting Future Demand

The MGSR trained on all available data was used to assess how changes in service capacity and population health might impact ED attendances in the future, compared to a ‘do-nothing’ scenario where the only independent variable that changes is population. Using the model to forecast ED attendances over a 4-year period, we found that changes to overall population health would have the largest instant impact (Figure 5). If population health in CCGs that were below average at baseline were improved year-on-year until the baseline average for each component was met, average peak ED attendances per CCG would be approximately 600 lower after 2 years (grey line, Figure 5) compared to a ‘do-nothing’ scenario (green dashed line, Figure 5). Increasing GP capacity by 10% would have no impact on mean ED attendances (purple line, Figure 5). Increasing Ambulance and 111 capacity would lead to an overall increase in ED attendances (blue and red lines, Figure 5). An increase in Ambulance capacity would have the greatest impact resulting in average peak ED attendances per CCG increasing by 200 per month.

**Figure 5:**
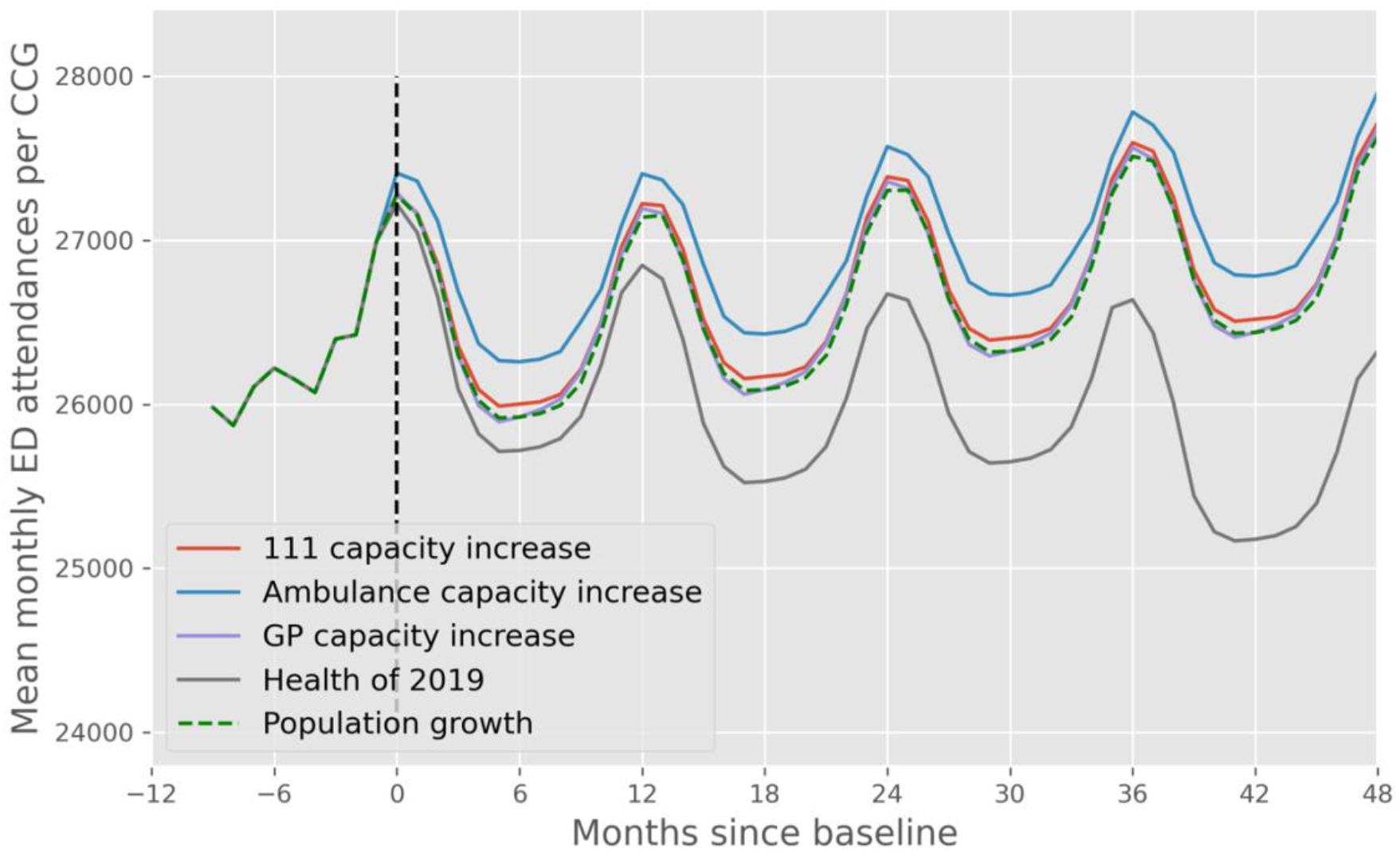
Mean monthly ED attendances per CCG forecasted over a 4-year period under 5 different scenarios. Scenarios: ‘do-nothing’ (green dashed line); increase 111 capacity by 10% (red line); increase ambulance capacity by 10% (blue line); increase GP capacity by 10% (purple line); if population health measures are less than the baseline average, increase them by 0.2 points per year until they reach the baseline average (grey line). The black dashed line represents the transition from baseline values (not predicted) to values predicted using the MGSR. All lines correspond to a 4-month rolling average of the monthly values.

## Discussion

We have presented a novel MGSR model that can be used to forecast monthly ED attendances for CCGs in England. The MGSR combines measures of health service capacity (Capacity model) and measures of population health (Population Health model). The Capacity model can explain 41 ± 4% of the variance in monthly ED attendances, and the Population Health model explains 61 ± 25% of the variance. Combining these two models, the MGSR has an overall R-squared (variance explained) of 0.75 ± 0.03% which is an improvement on the performance of each of the component models independently. Alongside an increase in R-squared, the MGSR allows for monthly ED attendances to be forecast while accounting for annual changes in population size and measures of population health. The MGSR trained on data from 2018 was able to predict mean monthly ED attendances per CCG in 2019 with, on average, only a 4% error. Using the MGSR to forecast future ED attendances we found that improving population health would lead to a reduction in mean monthly ED attendances however changes to service capacity may lead to an increase.

When using the MGSR to forecast future ED demand, individual predictions of monthly ED attendances for each CCG were aggregated into an average across CCGs. As such, it is important to note that while the model suggests improving population health will result in a reduction in ED attendances, and increasing service capacity may increase ED attendances, these relationships only hold at the aggregate level. For an individual CCG, the relationship between population health, service capacity and monthly ED attendances can be different to that which is seen globally [35].

A strength of the MGSR model is that it was developed using publicly available data from 2018-2019 aggregated to CCG regions. We selected this study period due to the impact of COVID-19 on health service capacity and service accessibility; a model trained on data from 2020 and 2021 would not be transferable to the post-Covid era due to health service disruption during the pandemic. As the data we have used is updated either monthly [26,27,29] or annually [28,30,32], once enough data is available from the post-Covid era, the model can be re-trained to produce more up-to-date forecasts without changing any of the underlying architecture. While recent changes in the organisational structure of the NHS mean that CCGs no longer exist, having been replaced with Integrated Care Boards (ICBs), any future data can be mapped to past CCG regions (now referred to as sub-ICBs) or the model can be re-trained at the geographical level of ICBs.

We used RFs for the Population Health and Capacity models as they performed best when compared to linear models and other ensembles [35]. While we have compared multiple ML and statistical approaches, the list of models implemented was not exhaustive. Previous work has demonstrated that Neural Networks (NNs) are superior to linear models for predicting short-term ED attendances in certain settings [5,14], and time-series methods, such as ARIMAX, can outperform ML approaches [8,10]. We did not consider these approaches due to data availability; both NNs and ARIMAX models require a large amount of data during training and ARMIAX would not be suitable for long-term forecasts when the values of the exogenous (independent) variables are not known, as is the case for the variables used here. Furthermore, while NNs often outperform other ML approaches, this is at the cost of interpretability. To ensure our model is useful in practice, in the RFs we selected the number and size of trees that optimised performance while minimising complexity [35].

One limitation of RFs is that they are not able to make predictions outside of the range of the data seen during training. This can be seen in Figure 2 where the range of predicted values (y-axis) is smaller than the range of true values (x-axis). Consequently, the MGSR will not perform as well in regions with a very low or very high number of ED attendances per 10,000 people.

Comparing the monthly ED attendances predicted by the MGSR to their true values (Figure 2) it is clear that the MGSR does not always perform well. Specifically, from this figure the two clusters of points above the dashed line appear to be outliers. Further analysis found that each cluster represents a distinct CCG. These two CCGs have very low values for *People* and *Lives* [35]. As the population health model contains variables that vary annually and our study period is two years, the amount of data available for training this model was limited to 145 data points. This smaller sample of data resulted in less accurate predictions for CCGs that had very low or high values for any of the population health variables. This limitation will be overcome in the future once more data becomes available.

When using the MGSR to forecast future ED demand, improving population health was the only scenario that resulted in a decrease in ED attendances (Figure 5). Of the population health variables, *Lives*, which quantifies health-related behaviours and personal circumstances, was the most important (PI 0.20 ± 0.04, Figure 4). For policy makers, this suggests that interventions to reduce the prevalence of smoking and alcohol abuse, tackle obesity and improve vaccination coverage could all have an impact on long term ED demand. As each of the domains of the Health Index (*People, Places* and *Lives*) are an aggregate of underlying components, future work looking at the importance of each sub-component and how changes in these impact ED demand would be beneficial to policy makers: knowledge of the underlying component(s) that have the greatest impact on ED attendances would ensure policies are targeted and effective.

In contrast to improving population health, increasing ambulance capacity led to an overall increase in ED attendances (Figure 5). This may be explained by the fact that monthly ambulance capacity is lower than utility for all CCGs: the number of calls answered is always less than the number of calls made [29,35]. Therefore, if ambulance capacity is increased, meaning more calls are answered, more people will be conveyed to hospital. The model is not able to predict what would happen in a situation where no data are available, for example where ambulance capacity is greater than utility. While this is clearly a limitation of the model, it is also a limitation of the data and the way the ambulance service in England currently operates: if the ambulance service is always saturated, it is not possible to use data to model a scenario when it is unsaturated.

If the model were to be used locally by, e.g. an ICB, it would significantly benefit from being re-trained with local data at a lower level of geography. This would ensure that the model is tailored to the local population. For example, if the independent variables were calculated per Lower Super Output Area (LSOA) the model could be used to forecast total monthly ED attendances within an Integrated Care System (ICS) under different practical scenarios. Using local data would also help to overcome the limitation of the model not performing well for regions that are outliers in terms of population health; ICS regions cover on average 1.5 million people [37] equating to approximately 850 LSOAs per ICS (or 1700 values of population health variables over two years) compared to the 74 CCGs used in this study.

Our MGSR model can be used by both strategic commissioners and policy makers. For commissioners, the Capacity model can be used to assess how additional spending, or spending cuts, in other services will impact monthly ED attendances. Using the model to determine how spending on one service impacts other services would allow for both cost and service capacity to be optimised across a region. The Population Health model can be used by policy makers to understand how health interventions might impact ED attendances. Working alongside service planners, the value of an intervention could be assessed through its impact on both population health and reduction in cost for the local health service.

## Conclusions

To our knowledge this is the first model that combines measures of wider urgent care service capacity and population health for forecasting long-term demand on ED departments. We demonstrated that 75% of the variance in monthly ED attendances can be explained using only 7 variables obtained from publicly available data. Using the MGSR to forecast mean monthly ED attendances for 74 CCGs in the UK, we found that improving population health would have the largest impact. Our model can be used by strategic planners and policy makers to measure the impact of future changes or interventions on safety and quality of services.

## Data Availability

All data produced are available online at https://github.com/CharlotteJames/ed-forecast/tree/main/data

https://charlottejames.github.io/ed-forecast/

https://www.england.nhs.uk/statistics/statistical-work-areas/ae-waiting-times-and-activity/

https://digital.nhs.uk/data-and-information/publications/statistical/appointments-in-general-practice

https://www.england.nhs.uk/statistics/statistical-work-areas/iucadc-new-from-april-2021/nhs-111-minimum-data-set

https://www.england.nhs.uk/statistics/statistical-work-areas/ambulance-quality-indicators

https://www.ons.gov.uk/peoplepopulationandcommunity/populationandmigration/populationestimates/datasets/clinicalcommissioninggroupmidyearpopulationestimates

https://www.ons.gov.uk/peoplepopulationandcommunity/healthandsocialcare/healthandwellbeing/datasets/healthindexscoresengland

## List of Abbreviations

ED: Emergency Department
MGSR: Multi-Granular Stacked Regression
MAPE: Mean Absolute Percentage Error
ARIMA: Auto-Regressive Integrated Moving Average
ML: Machine Learning
GP: General Practice
CCG: Clinical Commissioning Group
ONS: Office for National Statistics
RF: Random Forest
OLS: Ordinary Least Squares
GI: Gini Importance
PI: Permutation Importance
ICB: Integrated Care Board
NN: Neural Network
LSOA: Lower Super Output Area
ICS: Integrated Care system

## Declarations

### Ethics approval and consent to participate

Not applicable

### Consent for publication

Not applicable

### Availability of data and materials

The data generated and analysed during the study are available on GitHub: https://github.com/CharlotteJames/ed-forecast/tree/main/data

### Competing interests

All authors declare no competing interests

### Funding

CJ and RD are funded by NIHR Bristol BRC (IS_BRC_1215_20011). CJ is funded by NIHR Research Capability Funding (RCF 21/22-4.2). RD is funded by HDR UK South West CFC0129.

### Authors’ contributions

All authors contributed to the design of the study. CJ acquired the data, carried out the modelling and analysis and drafted the manuscript. All authors contributed to the final version of the manuscript.

## Acknowledgements

Not applicable

## Notes

### Competing Interest Statement

The authors have declared no competing interest.

### Funding Statement

This study was funded by NIHR Bristol BRC, NIHR Research Capability Funding and HDR UK South West

### Author Declarations

The study used only openly available data that were originally located at: https://www.ons.gov.uk/peoplepopulationandcommunity/populationandmigration/populationestimates/datasets/; https://www.england.nhs.uk/statistics/statistical-work-areas; https://www.ons.gov.uk/peoplepopulationandcommunity/healthandsocialcare/healthandwellbeing/datasets/; https://digital.nhs.uk/data-and-information/publications/statistical/

## References

[1] The Kings Fund, What’s going on with A&E waiting times? 2022. https://www.kingsfund.org.uk/projects/urgent-emergency-care/urgent-and-emergency-care-mythbusters Accessed 18 August 2022

[2] House of Commons Library, NHS Key Statistics: England, February 2020. 2020. https://researchbriefings.files.parliament.uk/documents/CBP-7281/CBP07281-feb2020.pdf Accessed 18 August 2022

[3] Turner J, Knowles E, Simpson R, Sampson F, Dixon S, Long J, Bell-Gorrod H, Jacques R, Coster J, Yang H, Nicholl J. Impact of NHS 111 Online on the NHS 111 telephone service and urgent care system: a mixed-methods study. Health Services and Delivery Research. 2021;9(21):1–48.

[4] Cheng Q, Argon NT, Evans CS, Liu Y, Platts-Mills TF, Ziya S. Forecasting emergency department hourly occupancy using time series analysis. Am J Emerg Med. 2021 Oct;48:177–82.

[5] Zhang Y, Zhang J, Tao M, Shu J, Zhu D. Forecasting patient arrivals at emergency department using calendar and meteorological information. Appl Intell (Dordr). 2022 Jan 21;1–12.

[6] Boyle J, Jessup M, Crilly J, Green D, Lind J, Wallis M, et al. Predicting emergency department admissions. Emerg Med J. 2012 May;29(5):358–65.

[7] Ordu, M., Demir, E., & Tofallis, C. (2019). A comprehensive modelling framework to forecast the demand for all hospital services. The International Journal of Health Planning and Management. DOI: 10.1002/hpm.2771

[8] Jones SS, Thomas A, Evans RS, Welch SJ, Haug PJ, Snow GL. Forecasting daily patient volumes in the emergency department. Acad Emerg Med. 2008 Feb;15(2):159–70

[9] Tideman S, Santillana M, Bickel J, Reis B. Internet search query data improve forecasts of daily emergency department volume. J Am Med Inform Assoc. 2019 Dec 1;26(12):1574–83.

[10] Vollmer MAC, Glampson B, Mellan T, Mishra S, Mercuri L, Costello C, et al. A unified machine learning approach to time series forecasting applied to demand at emergency departments. BMC Emerg Med. 2021 Jan 18;21(1):9.

[11] Champion R, Kinsman LD, Lee GA, Masman KA, May EA, Mills TM, et al. Forecasting emergency department presentations. Aust Health Rev. 2007 Feb;31(1):83–90.

[12] Jilani T, Housley G, Figueredo G, Tang PS, Hatton J, Shaw D. Short and Long term predictions of Hospital emergency department attendances. Int J Med Inform. 2019 Sep;129:167–74.

[13] Zhou L, Zhao P, Wu D, Cheng C, Huang H. Time series model for forecasting the number of new admission inpatients. BMC Med Inform Decis Mak. 2018 Jun 15;18(1):39.

[14] Sudarshan VK, Brabrand M, Range TM, Wiil UK. Performance evaluation of Emergency Department patient arrivals forecasting models by including meteorological and calendar information: A comparative study. Comput Biol Med. 2021 Aug;135:104541.

[15] Scantlebury R, Rowlands G, Durbaba S, Schofield P, Sidhu K, Ashworth M. Socioeconomic deprivation and accident and emergency attendances: cross-sectional analysis of general practices in England. Br J Gen Pract. 2015 Oct;65(639):e649–654.

[16] Tuominen J, Lomio F, Oksala N, Palomäki A, Peltonen J, Huttunen H, Roine A. Forecasting daily emergency department arrivals using high-dimensional multivariate data: a feature selection approach. BMC Med Inform Decis Mak. 2022 Dec;22(1):1–2.

[17] Giebel C, McIntyre JC, Daras K, Gabbay M, Downing J, Pirmohamed M, et al. What are the social predictors of accident and emergency attendance in disadvantaged neighbourhoods? Results from a cross-sectional household health survey in the north west of England. BMJ Open. 2019 Jan 6;9(1):e022820.

[18] Hull SA, Homer K, Boomla K, Robson J, Ashworth M. Population and patient factors affecting emergency department attendance in London: retrospective cohort analysis of linked primary and secondary care records. Br J Gen Pract. 2018 Mar;68(668):e157–67.

[19] Scantlebury R, Rowlands G, Durbaba S, Schofield P, Sidhu K, Ashworth M. Socioeconomic deprivation and accident and emergency attendances: cross-sectional analysis of general practices in England. Br J Gen Pract. 2015 Oct;65(639):e649–654.

[20] Pedrycz W. Granular computing: an introduction. InProceedings joint 9th IFSA world congress and 20th NAFIPS international conference (Cat. No. 01TH8569) 2001 Jul 25 (Vol. 3, pp. 1349-1354). IEEE.

[21] Kumar N, Baghel BK. Smart stacking of deep learning models for granular joint intent-slot extraction for multi-intent SLU. IEEE Access. 2021 Jul 7;9:97582–90.

[22] Jin J, Zhao Y, Cui R. Research on Multi-granularity Ensemble Learning Based on Korean. In The 2nd International Conference on Computing and Data Science 2021 Jan 28 (pp. 1-6).

[23] Zhao Q, Lyu S, Li Y, Ma Y, Chen L. MGML: Multi-Granularity Multi-Level Feature Ensemble Network for Remote Sensing Scene Classification. IEEE Trans Neural Netw Learning Syst. Published online 2021:1-15. Doi:10.1109/TNNLS.2021.3106391

[24] Deng W, Wang G, Zhang X, Xu J, Li G. A multi-granularity combined prediction model based on fuzzy trend forecasting and particle swarm techniques. Neurocomputing. 2016 Jan 15;173:1671–82.

[25] Wang J, Zhou H, Hong T, Li X, Wang S. A multi-granularity heterogeneous combination approach to crude oil price forecasting. Energy Economics. 2020 Sep 1;91:104790

[26] NHS England. A&E Attendances and Emergency Admissions. 2018-2019. https://www.england.nhs.uk/statistics/statistical-work-areas/ae-waiting-times-and-activity/ Accessed 18 August 2022

[27] NHS Digital. Appointments in General Practice. 2018-2019. https://digital.nhs.uk/data-and-information/publications/statistical/appointments-in-general-practice Accessed 18 August 2022

[28] NHS England. NHS 111 Minimum Dataset. 2018-2019. https://www.england.nhs.uk/statistics/statistical-work-areas/iucadc-new-from-april-2021/nhs-111-minimum-data-set Accessed 18 August 2022

[29] NHS England. Ambulance Quality Indicators. 2018-2019. https://www.england.nhs.uk/statistics/statistical-work-areas/ambulance-quality-indicators Accessed 18 August 2022

[30] Office for National Statistics. Clinical Commissioning Group Mid-year Population Estimates. 2018. https://www.ons.gov.uk/peoplepopulationandcommunity/populationandmigration/populationestimates/datasets/clinicalcommissioninggroupmidyearpopulationestimates Accessed 18 August 2022

[31] Office for National Statistics. Developing the Health Index for England. 2020. https://www.ons.gov.uk/peoplepopulationandcommunity/healthandsocialcare/healthandwellbeing/articles/developingthehealthindexforengland/2015to2018 Accessed 18 August 2022

[32] Office for National Statistics. Health Index Scores, England. 2022 https://www.ons.gov.uk/peoplepopulationandcommunity/healthandsocialcare/healthandwellbeing/datasets/healthindexscoresengland Accessed 18 August 2022

[33] Breiman L. Random forests. Machine learning. 2001 Oct;45(1):5–32.

[34] Pedregosa F, Varoquaux G, Gramfort A, Michel V, Thirion B, Grisel O, Blondel M, Prettenhofer P, Weiss R, Dubourg V, Vanderplas J. Scikit-learn: Machine learning in Python. the Journal of machine Learning research. 2011 Nov 1;12:2825–30.

[35] James C. Forecasting Long Term Emergency Department Demand. 2022. https://charlottejames.github.io/ed-forecast/ doi: 10.5281/zenodo.7157034 Accessed 18 August 2022

[36] Hastie T, Tibshirani R, Friedman JH, Friedman JH. The elements of statistical learning: data mining, inference, and prediction. New York: springer; 2009 Aug.

[37] NHS England. Allocation of Resources 2022/23. 2022. https://www.england.nhs.uk/wp-content/uploads/2022/04/integrated-care-board-allocation-core-services.xlsx Accessed 18 August 2022

